# Causal Relationship between Mental Disorders and Cancers: a Mendelian Randomization Study

**DOI:** 10.1101/2024.07.23.24310860

**Authors:** Bowen Du, Han Hong, Chaopeng Tang, Li Fan, Jie Dong, Jingping Ge, Xuejun Shang

**Affiliations:** Department of Urology, Jinling Hospital, Affiliated Hospital of Medical School, Nanjing University, Nanjing, China

**Keywords:** Mental disorders, Cancer, Mendelian randomization, Genetics, SNP

## Abstract

**Background:** Evidence from observational studies suggests an association between mental disorders and cancers. However, the causality of this association remains unclear.

**Methods:** We collected genome-wide association study (GWAS) summary statistics of five mental disorders from the Psychiatric Genomics Consortium (PGC, 72,517 to 500,199 participants), paired with GWAS summary statistics of the risks of 18 cancer types from the UK Biobank (167,020 to 361,194 participants) and FinnGen database (110,521 to 264,701 participants). We conducted univariable and multivariable Mendelian randomization (MR) analyses to explore the causal relationships.

**Results:** We identified ten causal associations between mental disorders and cancer risks. Notably, anorexia nervosa (AN) exhibits a causal association with a decreased risk of prostate cancer (β = -0.30, p = 1.61 × 10^-6^) and an elevated risk for stomach cancer (β = 0.47, p = 5.3 × 10^-3^). Bipolar disorder (BD) is causally linked to a reduced risk of pancreatic cancer (β = -5.13 × 10^-4^, p = 3.2 × 10^-3^). Major depression disorder (MDD) is causally associated with an elevated risk of bladder cancer (β = 1.84 × 10^-3^, p = 5.0 × 10^-4^) and kidney cancer (β = 1.40 × 10^-3^, p = 4.9 × 10^-3^). Additionally, we found the causal effect of skin melanoma on BD (β = -10.39, p = 2.1×10^-4^) and Schizophrenia (SCZ, β = -7.42, p = 3.3 × 10^-4^) with a bi-directional MR analysis. Moreover, we identified leukocyte count as a causal mediator of a causal association between AN and stomach cancer with a two-step MR analysis.

**Conclusions:** In summary, our MR analysis reveals that mental disorders were causally associated with cancer risks.

## Background

Mental disorders, such as attention-deficit and hyperactivity disorder (ADHD), anorexia nervosa (AN), bipolar disorder (BD), major depression disorder (MDD), and schizophrenia (SCZ), pose a considerable global burden on human health [1]. Meanwhile, cancer also represents a significant public health challenge worldwide [2]. Existing observational studies have established associations between mental disorders and various types of cancers. For example, ADHD is correlated with an increased risk of colorectal cancer [3], while AN is implicated in elevated risk for both lung cancer and esophageal cancer [4], as well as a reduced risk of breast cancer [5]. BD appears to be associated with an increased risk of multiple types of cancers, particularly breast cancer [6, 7]. Moreover, individuals with depressive symptoms experience higher mortality rates and diminished disease-free survival in the context of bladder cancer [8]. SCZ is associated with an increased risk of breast cancer [9] but exhibits a decreased risk for colorectal and prostate cancer [10, 11].

While evidence indicates a correlation between mental disorders and various cancers, the causality underlying these associations has not been fully investigated, primarily due to unmeasured confounding variables or reverse causation. Advances in large-scale genome-wide association study (GWAS) and the methodology of Mendelian randomization (MR) have enabled researchers to investigate the causal relationships between risk factors and disease outcomes. Existing studies investigating the causal relationships between mental disorders and cancers have predominantly focused on breast cancer [12–14]. Nevertheless, comprehensive investigations into the causal association between mental disorders and a broader range of cancers are still lacking. Moreover, the mechanisms by which mental disorders influence cancer risks also need to be investigated.

In this study, we conducted a comprehensive two-sample univariable MR (UVMR) analysis to explore causal relationships between five distinct mental disorders and 18 types of cancer. Additionally, employing a bi-directional MR approach, we assessed the potential causal effect of cancers on mental disorders. To ascertain the reliability of our findings, we performed a multivariable MR (MVMR) analysis. Finally, we sought to identify potential mediators of the causal associations, utilizing a two-step MR approach.

## Methods

### Datasets

All analyses in this investigation relied on summary-level GWAS data. We sourced the summary statistics for GWAS of mental disorders from the Psychiatric Genomics Consortium (PGC) - a large-scale, multi-ethnic psychiatric GWAS repository composed of over 400,000 participants. We examined a total of five mental disorders: attention deficit hyperactivity disorder (ADHD), bipolar disorder (BD), anorexia nervosa (AN), major depressive disorder (MDD), and schizophrenia (SCZ).

We employed 54 GWAS summary statistics encompassing 18 types of cancers, which were sourced from the UK Biobank, composed of ∼500,000 participants [15], and FinnGen datasets [16], composed of ∼370,000 participants, as our outcome data. The UK Biobank GWAS summary statistics were acquired from the Neale Lab, while the FinnGen GWAS summary statistics were downloaded from the FinnGen website. We utilized GWAS summary statistics for 65 clinical traits from the UK Biobank dataset as potential mediators in the two-step MR analysis. These summary statistics were acquired from the Neale Lab. All the GWAS data used in this study were restricted to European ancestry. Comprehensive details of all GWAS summaries are provided in Table S1.

### Instrumental variable selection

The genetic instrument variables (IVs), typically single-nucleotide polymorphisms (SNPs), were derived from GWAS summary statistics utilizing the TwoSampleMR R package. For IV clumping, the p-value was set to 5×10^-8^; other parameters were set with R^2^ < 0.01 and clumping distance = 250 kb. In the data harmonization process, both palindromic and duplicated SNPs were excluded.

Instrumental variable (IV) outliers were identified using the MR-PRESSO R package. For each SNP, the MR-PRESSO outlier test calculated a p-value to assess its significance for pleiotropy, while the MR-PRESSO global test calculated a p-value for the overall presence of horizontal pleiotropy. SNPs with a p-value less than 0.05 were removed if the global p-value was less than 0.05. Subsequently, the global p-value was recalculated to evaluate the extent of pleiotropy.

### UVMR analysis

We conducted a two-sample UVMR analysis to investigate the causal relationship between mental disorders and cancers, employing the TwoSampleMR R package. We utilized five established MR methods for MR analysis with multiple IVs: MR Egger (MRE), weighted median (WM), inverse variance weighted (IVW), Simple mode, and Weighted mode. Analyses with fewer than three IVs were excluded, as an inadequate number of IVs would compromise the reliability of the analysis. Furthermore, we established a multiple testing significance threshold: p < 0.05/n, where n is the number of exposures [17].

To mitigate biases introduced by weak IVs, we calculated the F-statistic according to the reference [18]. Initially, we computed the proportion of variance explained by each IV (PVE) using the equation [19]:

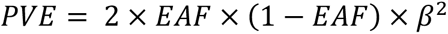

where EAF represents the effect allele frequency (EAF), and β is the effect size. The F-statistic is subsequently calculated using the formulation:

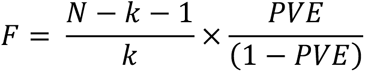

where N is the sample size and k is the number of IVs. We computed the mean F-statistic across all IVs. The F-statistic for AN was not calculated because the EAF information is not available according to the policies of the PCG. Additionally, the MR Steiger directionality test was used to examine whether exposure was directionally causal for the outcome [20].

### Heterogeneity and pleiotropy test

We evaluated the heterogeneity of MR analysis using Cochran’s Q test, designating a p-value (P_Q_) less than 0.05 as the existence of heterogeneity. Pleiotropy was assessed via MR-Egger regression [21] and MR-PRESSO residuals tests, considering a p-value (P_Egger_ or P_PRESSO_) less than 0.05 as evidence of pleiotropy. P_Q_ and P_Egger_ were calculated using the TwoSampleMR R package. P_PRESSO_ was computed with the MRPRESSO R package. To ensure the reliability of our findings, we excluded the analyses with P_Q_, P_Egger_, or P_PRESSO_ below 0.05 from the results.

### Bi-directional MR analysis

To investigate the causal effect of cancers on mental disorders, we conducted a bi-directional MR analysis, using cancer risks as exposures and mental disorders as outcomes. The bi-directional MR analysis was executed in accordance with the procedure of UVMR analysis.

### MVMR analysis

Considering potential genetic correlations among exposures may introduce inaccuracies in UVMR analysis [22], we employed MVMR analysis using the MVMR-IVW method. Since mental disorders are associated with smoking, alcohol consumption, and insomnia [23–25], the analyses were adjusted for the confounding variables, including smoking, alcohol consumption, and insomnia within the European population. The MVMR analysis was executed using the TwoSampleMR R package and was conducted separately for each mental disorder. The IVs were selected based on the following criteria: a p-value threshold of 5×10^-8^, R^2^ less than 0.01, and a clumping distance of 250 kb.

### Two-step MR analysis

The two-step MR analysis was conducted to elucidate mediators in the causal association between mental disorders and cancer risks. We selected 65 clinical traits as the potential mediators. To avoid the overlap of samples, we used the GWAS summary statistics of mental disorders from PGC as the exposures, the GWAS summary statistics of clinical traits from the UK biobank as the mediators, and the GWAS summary statistics of cancer risks from FinnGen as the outcomes.

The two-step MR analysis was performed according to the guidelines [26, 27]. First, the causal effect of mental disorders on clinical traits (β_1_) was evaluated with UVMR analysis. Second, the causal effect of clinical traits on cancers (β_2_) was measured with UVMR analysis. We used separate IVs for mental disorders and clinical traits. The total causal effect of mental disorders on cancers (β_0_) was measured above. The indirect causal effect was measured with β_1_ × β_2_. The direct causal effect was measured with β_0_ - β_1_ × β_2_. The proportion of indirect causal effect was measured with (β_1_ × β_2_) / β_0_.

## Results

### Study design

The design of this study is presented in Fig. 1. First, we applied UVMR analysis to investigate the causal associations between mental disorders and cancer risks. Second, we employed a bi-directional MR analysis to explore the causal effects of cancers on mental disorders. Third, we evaluated the reliability of causal associations with an MVMR analysis. Last, we used a two-step MR analysis to identify the mediators for the causal associations between mental disorders and cancers. The details of all GWAS summaries are presented in Table S1.

**Figure 1.**
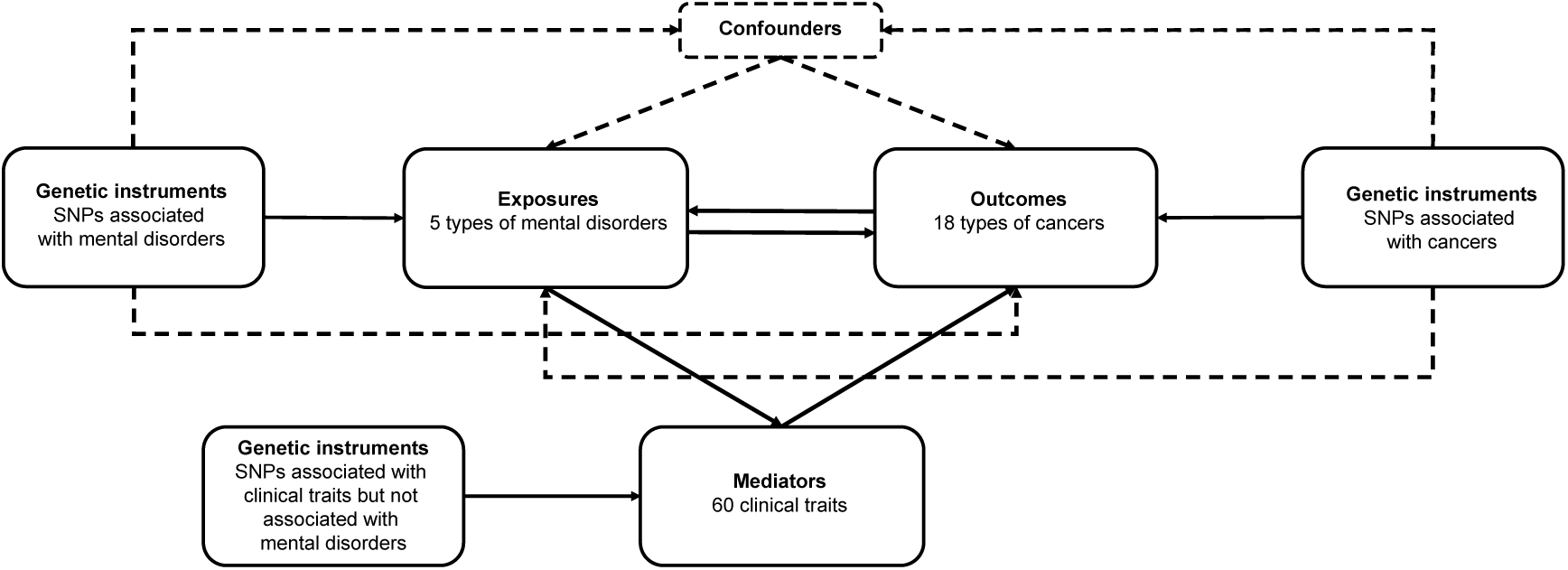
Design of this study. Univariable MR analysis was applied to investigate the causal associations between mental disorders and cancer risks. The bi-directional MR analysis was used to explore the causal effects of cancers on mental disorders. Two-step MR analysis was applied to identify the mediators for the causal associations between mental disorders and cancer risks. Three assumptions should be kept in MR analysis: (1) genetic variants must be associated with exposures, (2) genetic variants should not be associated with confounders, and (3) genetic variants must affect outcomes only through exposures, not through other pathways. Five types of mental disorders include attention deficit hyperactivity disorder (ADHD), anorexia nervosa (AN), bipolar disorder (BD), major depressive disorder (MDD), and schizophrenia (SCZ). Eighteen types of cancer include bladder cancer (BLCA), brain cancer (BNCA), brain cancer (BRCA), colon cancer (COCA), corpus uteri cancer (CPCA), esophageal cancer (ESCA), kidney cancer (KICA), liver cancer (LICA), lung cancer (LUCA), mesothelioma (MESO), ovarian cancer (OVCA), pancreatic cancer (PACA), prostate cancer (PRCA), rectal cancer (RECA), skin melanoma (SKCM), stomach cancer (STCA), testicular cancer (TECA), and thyroid cancer (THCA). Sixty clinical traits were listed in Supplementary Table 1.

### Causal effects of mental disorders on cancers

We investigated the causal effect of five specific mental disorders (ADHD, AN, BD, MDD, and SCZ) on 18 distinct types of cancers with UVMR analysis. We utilized five established MR methods for MR analysis: MR Egger (MRE), weighted median (WM), inverse variance weighted (IVW), Simple mode, and Weighted mode. The results are presented in Table 1 and Table S2. We identified ten causal associations between mental disorders and cancer risks, referring to AN, BD, MDD, and SCZ. No significant causal association with ADHD was observed.

**Table 1.** MR results of causal effects between mental disorders and cancers.

| Mental disorder | Cancer | Phenotype code | Dataset | Method | Number of SNP | $\beta$ | SE of $\beta$ | p value | OR (0.95 CI) |
| --- | --- | --- | --- | --- | --- | --- | --- | --- | --- |
| AN | Colon cancer | C18 | UK Biobank | Weighted median | 8 | $-3.33 \times 10^{-3}$ | $1.09 \times 10^{-3}$ | $2.35 \times 10^{-3}$ | 0.997 (0.995-0.999) |
| | Mesothelioma | C45 | UK Biobank | IVW | 8 | $-5.58 \times 10^{-4}$ | $2.07 \times 10^{-4}$ | $6.87 \times 10^{-3}$ | 0.999 (0.999-1.000) |
| | Pancreatic cancer | C25 | UK Biobank | Weighted median | 8 | $-1.34 \times 10^{-3}$ | $4.47 \times 10^{-4}$ | $2.75 \times 10^{-3}$ | 0.999 (0.998-1.000) |
| | Pancreatic cancer | C25 | UK Biobank | IVW | 8 | $-1.17 \times 10^{-3}$ | $3.50 \times 10^{-4}$ | $8.25 \times 10^{-4}$ | 0.999 (0.998-1.000) |
| | Prostate cancer | C3_PROSTATE | FinnGen | Weighted median | 8 | -0.332 | 0.080 | $3.51 \times 10^{-5}$ | 0.717 (0.613-0.840) |
| | Prostate cancer | C3_PROSTATE | FinnGen | IVW | 8 | -0.300 | 0.063 | $1.61 \times 10^{-6}$ | 0.741 (0.656-0.838) |
| | Skin melanoma | C3_MELANOMA_SKIN | UK Biobank | Weighted median | 8 | $-4.11 \times 10^{-3}$ | $1.13 \times 10^{-3}$ | $2.77 \times 10^{-4}$ | 0.996 (0.994-0.998) |
| | Skin melanoma | C3_MELANOMA_SKIN | UK Biobank | IVW | 8 | $-3.39 \times 10^{-3}$ | $8.48 \times 10^{-4}$ | $6.46 \times 10^{-5}$ | 0.997 (0.995-0.998) |
| | Skin melanoma | C3_MELANOMA_SKIN | FinnGen | Weighted median | 8 | 0.412 | 0.146 | $4.66 \times 10^{-3}$ | 1.509 (1.135-2.007) |
| | Skin melanoma | C3_MELANOMA_SKIN | FinnGen | IVW | 8 | 0.380 | 0.114 | $8.68 \times 10^{-4}$ | 1.462 (1.169-1.827) |
| | Stomach cancer | C3_STOMACH | FinnGen | Weighted median | 8 | 0.559 | 0.211 | $8.03 \times 10^{-3}$ | 1.749 (1.157-2.644) |
| | Stomach cancer | C3_STOMACH | FinnGen | IVW | 8 | 0.470 | 0.169 | $5.31 \times 10^{-3}$ | 1.600 (1.150-2.226) |
| BD | Pancreatic cancer | C3_PANCREAS | UK Biobank | Weighted median | 63 | $-7.49 \times 10^{-4}$ | $2.57 \times 10^{-4}$ | $3.57 \times 10^{-3}$ | 0.999 (0.999-1.000) |
| | Pancreatic cancer | C3_PANCREAS | UK Biobank | IVW | 63 | $-5.13 \times 10^{-4}$ | $1.74 \times 10^{-4}$ | $3.17 \times 10^{-3}$ | 0.999 (0.999-1.000) |
| | Pancreatic cancer | C25 | UK Biobank | Weighted median | 63 | $-7.69 \times 10^{-4}$ | $2.61 \times 10^{-4}$ | $3.26 \times 10^{-3}$ | 0.999 (0.999-1.000) |
| | Pancreatic cancer | C25 | UK Biobank | IVW | 63 | $-4.96 \times 10^{-4}$ | $1.80 \times 10^{-4}$ | $5.85 \times 10^{-3}$ | 1.000 (0.999-1.000) |
| MDD | Bladder cancer | C3_BLADDER | UK Biobank | IVW | 65 | $1.84 \times 10^{-3}$ | $5.30 \times 10^{-4}$ | $5.01 \times 10^{-4}$ | 1.002 (1.001-1.003) |
| | Bladder cancer | C67 | UK Biobank | IVW | 65 | $3.06 \times 10^{-3}$ | $7.55 \times 10^{-4}$ | $5.04 \times 10^{-5}$ | 1.003 (1.002-1.005) |
| | Kidney cancer | C64 | UK Biobank | IVW | 65 | $1.40 \times 10^{-3}$ | $4.97 \times 10^{-4}$ | $4.87 \times 10^{-3}$ | 1.001 (1.000-1.002) |
| SCZ | Pancreatic cancer | C3_PANCREAS | UK Biobank | MR Egger | 200 | $-7.82 \times 10^{-4}$ | $2.74 \times 10^{-4}$ | $4.85 \times 10^{-3}$ | 0.999 (0.999-1.000) |
| | Pancreatic cancer | C3_PANCREAS | UK Biobank | Weighted median | 200 | $-5.18 \times 10^{-4}$ | $1.47 \times 10^{-4}$ | $4.01 \times 10^{-4}$ | 0.999 (0.999-1.000) |
| | Pancreatic cancer | C3_PANCREAS | UK Biobank | IVW | 200 | $-3.05 \times 10^{-4}$ | $9.99 \times 10^{-5}$ | $2.26 \times 10^{-3}$ | 1.000 (0.999-1.000) |
| | Pancreatic cancer | C3_PANCREAS | UK Biobank | Weighted mode | 200 | $-8.13 \times 10^{-4}$ | $2.79 \times 10^{-4}$ | $3.97 \times 10^{-3}$ | 0.999 (0.999-1.000) |
| | Testis cancer | C3_TESTIS | UK Biobank | MR Egger | 200 | $-1.43 \times 10^{-3}$ | $5.48 \times 10^{-4}$ | $9.82 \times 10^{-3}$ | 0.999 (0.998-1.000) |
| | Testis cancer | C3_TESTIS | UK Biobank | IVW | 200 | $-5.21 \times 10^{-4}$ | $1.98 \times 10^{-4}$ | $8.54 \times 10^{-3}$ | 0.999 (0.999-1.000) |
| | Testis cancer | C3_TESTIS | UK Biobank | Weighted mode | 200 | $-1.61 \times 10^{-3}$ | $5.68 \times 10^{-4}$ | $4.99 \times 10^{-3}$ | 0.998 (0.997-1.000) |

AN, anorexia nervosa; BD, bipolar disorder; MDD, major depression disorder; SCZ, schizophrenia; IVW, inverse variance weighted

Our analyses demonstrate that AN exhibits a causal association with the decreased risk of several cancers: prostate cancer (β ± SE = -0.30 ± 0.06, p = 1.61 × 10^-6^, IVW), colon cancer [β ± SE = -(3.33 ± 1.09) × 10^-3^, p = 2.4 × 10^-3^, WM], mesothelioma [β ± SE = - (5.58 ± 2.07) × 10^-4^, p = 6.9 × 10^-3^, IVW], and pancreatic cancer [β ± SE = -(1.17 ± 0.35) × 10^-3^, p = 8.3 × 10^-4^, IVW]. These findings suggest a potential protective effect of AN on these specific types of cancers. Conversely, AN is causally associated with an elevated risk for stomach cancer (β ± SE = 0.47 ± 0.17, p = 5.3 × 10^-3^, IVW). Our analysis revealed conflicting results concerning the causal relationship between AN and skin melanoma across different databases. Specifically, AN was causally associated with a reduced risk of skin melanoma in the UK Biobank database [β ± SE = -(3.39 ± 0.85) × 10^-3^, p = 6.5 × 10^-5^, IVW], whereas it correlated with an elevated risk in the FinnGen database (β ± SE = 0.38 ± 0.11, p = 8.7 × 10^-4^, IVW). This contradictory result may be attributed to the population heterogeneity in these two databases.

Our findings suggest that BD is causally linked to a reduced risk of pancreatic cancer [β ± SE = -(5.13 ± 1.74) × 10^-4^, p = 3.2 × 10^-3^, IVW], potentially indicating a protective effect in the development of pancreatic cancer. Conversely, MDD is causally associated with an elevated risk of bladder cancer [β ± SE = (1.84 ± 0.53) × 10^-3^, p = 5.0 × 10^-4^, IVW] and kidney cancer [β ± SE = (1.40 ± 0.50) × 10^-3^, p = 4.9 × 10^-3^, IVW]. SCZ is causally associated with a reduced risk of pancreatic cancer [β ± SE = -(3.05 ± 1.00) × 10^-4^, p = 2.3 × 10^-3^, IVW] and testis cancer [β ± SE = -(5.21 ± 1.98) × 10^-4^, p = 8.5 × 10^-3^, IVW].

### Causal effects of cancers on mental disorders

To estimate the causal effect of cancers on mental disorders, we conducted a bi-directional MR analysis, using cancer risks as exposures and mental disorders as outcomes. Detailed results are delineated in Table 2 and Table S3. Specifically, skin melanoma is causally linked to a reduced risk of BD (β ± SE = -10.39 ± 2.80, p = 2.1 × 10^-4^, IVW) and SCZ (β ± SE = -7.42 ± 2.06, p = 3.3 × 10^-4^, IVW).

**Table 2.** Bi-directional MR analysis between cancers and mental disorders.

| Cancer | Phenotype code | Dataset | Mental disorder | Method | Number of SNP | $\beta$ | SE of $\beta$ | p value | OR (0.95 CI) |
| --- | --- | --- | --- | --- | --- | --- | --- | --- | --- |
| Skin melanoma | C3_MELANOMA_SKIN | UK Biobank | BD | WM | 10 | -12.01 | 2.91 | $3.70 \times 10^{-5}$ | $6.08 \times 10^{-6}$ ($2.02 \times 10^{-8}$ - $1.83 \times 10^{-3}$) |
| | C3_MELANOMA_SKIN | UK Biobank | BD | IVW | 10 | -10.39 | 2.80 | $2.09 \times 10^{-4}$ | $3.09 \times 10^{-5}$ ($1.27 \times 10^{-7}$ - $7.47 \times 10^{-3}$) |
| | C3_MELANOMA_SKIN | UK Biobank | SCZ | IVW | 10 | -7.42 | 2.06 | $3.25 \times 10^{-4}$ | $6.02 \times 10^{-4}$ ($1.06 \times 10^{-5}$ - $3.43 \times 10^{-2}$) |
| | C43 | UK Biobank | BD | IVW | 5 | -15.46 | 4.17 | $2.08 \times 10^{-4}$ | $1.93 \times 10^{-7}$ ($5.46 \times 10^{-11}$ - $6.82 \times 10^{-4}$) |
| | C3_MELANOMA_SKIN | FinnGen | SCZ | WM | 11 | -0.07 | 0.02 | $2.32 \times 10^{-4}$ | 0.934 (0.900 - 0.969) |
| | C3_MELANOMA_SKIN | FinnGen | SCZ | IVW | 11 | -0.06 | 0.02 | $2.71 \times 10^{-4}$ | 0.941 (0.911 - 0.972) |

BD, bipolar disorder; SCZ, schizophrenia; IVW, inverse variance weighted

### Causal effects independent of smoking, alcohol consumption, and insomnia

Prior research has established associations between mental disorders and lifestyle factors such as smoking, alcohol consumption, and insomnia [23–25]. To assess the influence of these covariates on causal relationships, we conducted an MVMR analysis using smoking, alcohol consumption, and insomnia as covariates. The findings are presented in Table 3 and Table S4. Referring to AN, BD, and MDD, the MVMR analysis confirms the causal association identified in the UVMR analysis (Table 3). Specifically, AN is causally associated with a decreased risk of prostate cancer (β ± SE = -0.20 ± 0.06, p = 0.030, IVW), colon cancer [β ± SE = -(1.58 ± 0.71) × 10^-3^, p = 0.026, IVW], mesothelioma [β ± SE = -(5.62 ± 1.82) × 10^-4^, p = 2.0 × 10^-3^, IVW], and pancreatic cancer [β ± SE = -(9.04 ± 3.39) × 10^-4^, p = 7.8 × 10^-3^, IVW]. Additionally, we identified a causal association between AN and an increased risk of stomach cancer (β ± SE = 0.31 ± 0.15, p = 0.042, IVW). BD is causally linked to a reduced risk of pancreatic cancer [β ± SE = -(4.56 ± 1.36) × 10^-4^, p = 7.8 × 10^-4^, IVW]. MDD is causally associated with an elevated risk of bladder cancer [β ± SE = (3.11 ± 0.65) × 10^-3^, p = 1.5 × 10^-6^, IVW] and kidney cancer [β ± SE = (1.46 ± 0.42) × 10^-3^, p = 4.8 × 10^-4^, IVW]. In contrast, SCZ shows no significant causal associations with either pancreatic cancer or testicular cancer when adjusted to those covariates. Nevertheless, our MVMR analyses affirm the robustness of our results concerning AN, BD, and MDD.

**Table 3.**
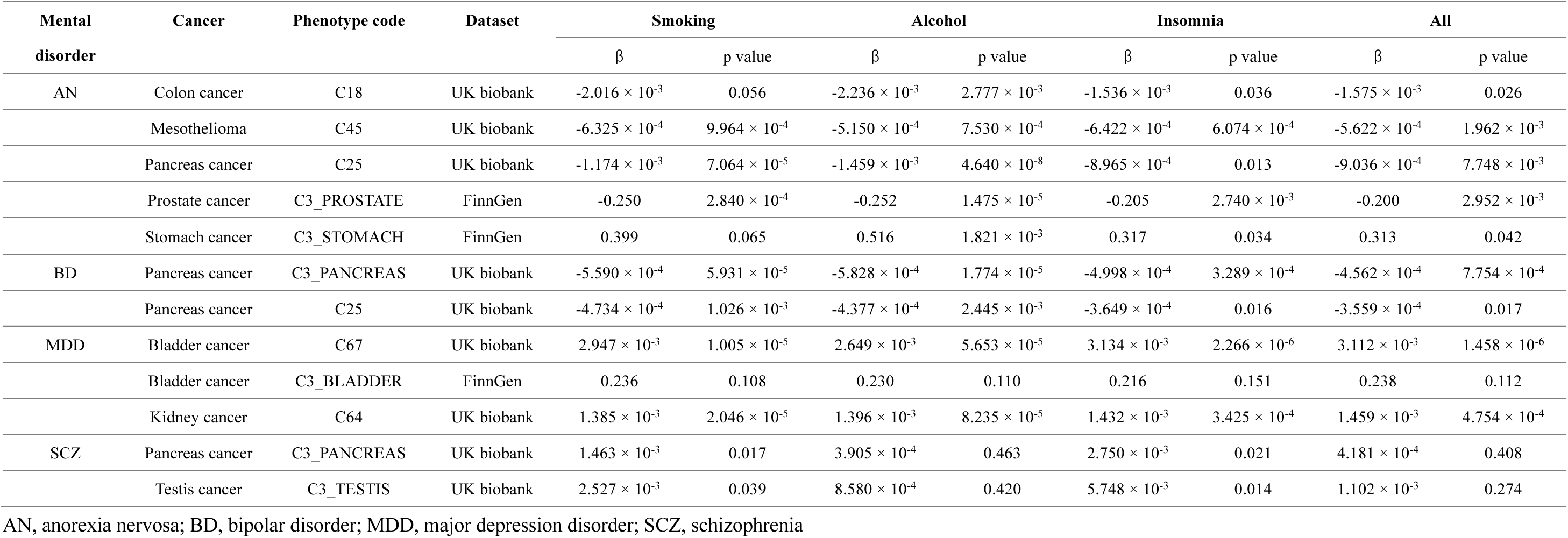
MVMR results of causal effects between mental disorders and cancers.

### The mediators of causal associations

Mental disorders may exert an influence on cancer development through intermediary factors, such as metabolites [28] and immune cells [29, 30]. To investigate the potential mediators, we employed a two-step MR analysis. This methodology comprised two steps: firstly, the assessment of the causal effect of five mental disorders on potential mediators, and secondly, the evaluation of the causal effect of these mediators on cancer risks. We selected 65 clinical traits as potential mediators, five of which were excluded due to insufficient IVs for analysis. The causal effects of mental disorders on clinical traits and the causal effects of clinical traits on cancer risks are individually delineated in Table S5 and Table S6.

As is shown in Fig. 2, leukocyte count serves as a mediator of the causal association between AN and stomach cancer. Specifically, AN is causally linked to a decreased leukocyte count (β ± SE = -0.048 ± 0.012, p = 4.7 × 10^-5^, IVW), whereas leukocyte count is causally associated with a decreased risk of stomach cancer (β ± SE = -1.336 ± 0.579, p = 0.022, Simple mode). The proportion of indirect causal effect is 13.6%. As low leukocyte count is an indicator of immune suppression [31], our finding suggests that AN promotes stomach cancer by inhibiting immune response.

**Figure 2.**
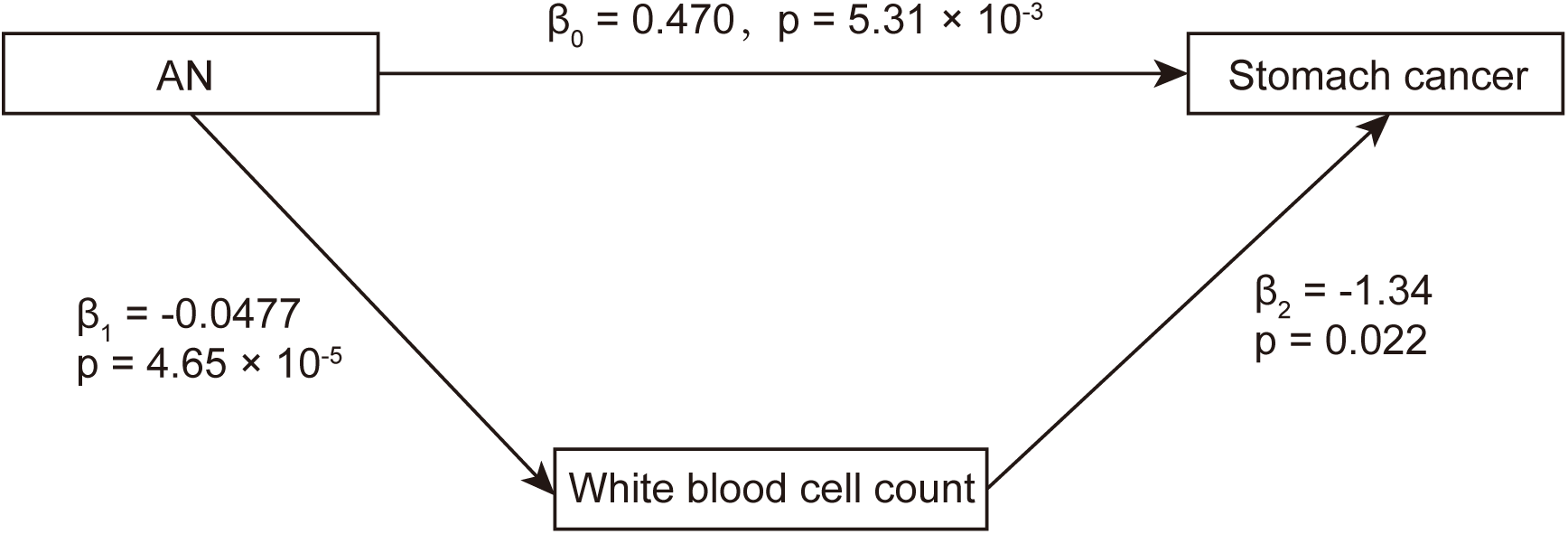
The summary of mediators of causal associations between mental disorders and cancers. Two-step MR analysis was used to identify the mediators of the causal associations. The schematic diagram shows the causal relationship between exposures (mental disorders), mediators (clinical traits), and outcomes (cancers). β_1_ indicates the causal effect of exposures on mediators. β_2_ indicates the causal effect of mediators on outcomes. β_0_ indicates the total causal effect of exposures on outcomes. The indirect causal effect of exposure on outcome is calculated by β_1_ multiplied by β_2_. The proportion of indirect effect is calculated using the following formula: proportion of indirect effect = (β_1_ × β_2_) / β_0_.

In addition to the results mentioned above, our two-step MR analysis yields insightful findings regarding the causal association between mental disorders and clinical traits. Specifically, BD, MDD, and SCZ are associated with increased levels of aspartate aminotransferase (BD: β ± SE = 0.031 ± 0.008, p = 8.4 × 10^-5^, IVW; MDD: β ± SE = 0.055 ± 0.016, p = 5.5 × 10^-4^, IVW; SCZ: β ± SE = 0.016 ± 0.004, p = 3.1 × 10^-4^, IVW; Fig. S1). MDD and SCZ are associated with increased levels of alanine aminotransferase (MDD: β ± SE = 0.094 ± 0.018, p = 1.8 × 10^-7^, WM; SCZ: β ± SE = 0.044 ± 0.014, p = 0.002, MRE; Fig. S1). This result indicates that these mental disorders may lead to liver injury. Moreover, our results reveal multiple potential carcinogenic factors. As is shown in Fig. S2, IGF-1 is associated with an increased risk of prostate cancer (β ± SE = 0.164 ± 0.037, p = 1.1 × 10^-5^, IVW) and breast cancer (β ± SE = 0.213 ± 0.053, p = 7.1 × 10^-5^, WM). Lymphocyte count is associated with an increased risk of pancreatic cancer (β ± SE = 0.753 ± 0.178, p = 2.9 × 10^-5^, MRE) and colon cancer (β ± SE = 0.707 ± 0.125, p = 2.7 × 10^-8^, MRE).

## Discussion

The investigation of causal relationships is substantially critical for elucidating the driving factors behind observed associations. In this study, we employed MR analyses to assess the causal links between mental disorders and cancers. Furthermore, we identified mediators of these causal associations, enhancing our understanding of how mental disorders influence cancer progression. To the best of our knowledge, this study, encompassing five mental disorders, 18 types of cancer, and 60 mediators, is the most comprehensive analysis to date on the causal relationship between mental disorders and cancers.

AN is associated with an increased risk of various types of cancers, including lung cancer and esophageal cancer [4, 32]. Our study demonstrates a causal link between AN and an elevated risk of stomach cancer. Our two-step MR analysis identified leukocyte count as a mediator in the causal relationship between AN and stomach cancer. AN is causally associated with a reduction in leukocyte count, which in turn is linked to a decreased risk of stomach cancer. As low leukocyte count is an indicator of a compromised immune system, AN may facilitate the development of stomach cancer by suppressing immune function. Our findings suggest that the dysregulation of immune cells plays a critical role in the causal associations between mental disorders and cancers.

There has been controversy over whether depression can lead to tumorigenesis. A prior study indicated that individuals with depression have an 18% heightened risk of developing cancer, notably lung cancer, gastrointestinal tract cancer, breast cancer, and urinary cancer [33]. Conversely, other research has found no significant correlation between depression and cancer incidence [34]. Our studies revealed that MDD is causally associated with an increased risk of bladder cancer. This study provides new evidence suggesting that depression may contribute to cancer development.

Prior research has suggested that mental disorders may correlate with a low incidence of cancer [4]. We found that several mental disorders are causally associated with a reduced risk of cancers. AN exhibits inhibitive causal effects on colon cancer, mesothelioma, pancreatic cancer, and prostate cancer. Moreover, BD is causally linked to a reduced risk of pancreatic cancer. These causal associations provide novel evidence for the notion that mental disorders may exhibit a protective effect on cancers in specific types of cancers.

Previous studies have reported a higher incidence of mental disorders in people with cancers [35, 36]. However, this finding is influenced by some factors such as clinical stage, cancer treatment, and so on [36]. Whether tumors can cause depression is far from a conclusion. Our bi-directional MR analysis indicates that skin melanoma is causally associated with a reduced risk of BD and SCZ, providing novel evidence for the notion that cancer may lead to mental disorders. Our research did not detect much causal effect of cancers on mental disorders, which may be attributed to these reasons: First, an insufficient number of IVs exists for MR analysis in many cancer types. Second, a stringent p-value threshold was applied to correct for multiple testing.

Nevertheless, this study has several limitations. First, our findings are derived from a European population, potentially leading to biased estimates and limited generalizability. Future research is required to investigate these causal relationships in diverse populations. Second, the strict criteria for multiple-testing correction may have resulted in the omission of some causal associations. Third, certain mental disorders, such as autism spectrum disorder, were excluded from this study due to the stringent p-value threshold for IVs. Fourth, while the F-statistic for AN is absent and the F-statistic for SCZ is below 10, this warrants caution regarding the weak IV bias for both AN and SCZ. Fifth, while we identified mediators of the causal association using a two-step MR analysis, experimental studies are still necessary to elucidate the mechanisms through which mental disorders affect cancer development.

## Conclusions

In summary, this study provides a comprehensive assessment of the causal relationship between mental disorders and diverse types of cancer using a two-sample MR analysis. Our results revealed multiple causal association between mental disorders and cancer risks. Additionally, our investigation demonstrated that leukocyte count may be a mediator of the causal associations between AN and stomach cancer. This study not only sheds light on the mechanisms of cancer development but also holds significant implications for the prevention and targeted therapy of cancers.

## Abbreviations

ADHD: Attention-deficit and hyperactivity disorder
AN: Anorexia nervosa
BD: Bipolar disorder
GWAS: Genome-wide association study
HDL: High-density lipoprotein
IV: Instrument variable
IVW: Inverse variance weighted
MDD: Major depression disorder
MR: Mendelian randomization
MRE: MR-Egger
MVMR: Mendelian randomization
SCZ: Schizophrenia
SM: Simple mode
SNP: Single-nucleotide polymorphisms
UVMR: Univariate Mendelian randomization
WM: weighted median

## Declarations

### Ethics approval and consent to participate

This study is performed using published studies and publicly available summary statistics. All original studies have been approved by the corresponding ethical review board. The participants have provided informed consent. Therefore, no new ethical review board approval was required.

### Consent for publication

Consent for publication was obtained from every individual whose data are included in this manuscript.

### Availability of data and material

The mental disorder GWAS summaries were downloaded from the PGC website (https://pgc.unc.edu/for-researchers/download-results/). The GWAS summary from UK Biobank were downloaded from the Neala Lab (http://www.nealelab.is/uk-biobank). The GWAS summary from FinnGen were downloaded following the instruction on its website (https://www.finngen.fi/en/access_results). No special codes were used in this study. The codes for MR analysis with the TwoSampleMR R package are available on the GitHub repository (https://github.com/BowenDuGroup/Mentaldisorderandcancer).

### Authors’ contributions

B.D. designed the study. B.D. and H.H. analyzed the data. C.T. , L.F., and J.D. curated and processed the data. B.D. wrote the manuscript. J.G. and X.S. supervised the research. All authors read and approved the final manuscript.

## Acknowledgements

The data analyzed in this study was provided by PGC consortium, UK Biobank, FinnGen and Neala Lab. We gratefully acknowledge their contributing studies and the participants in these studies.

## Supplementary Figures

**Figure S1.**
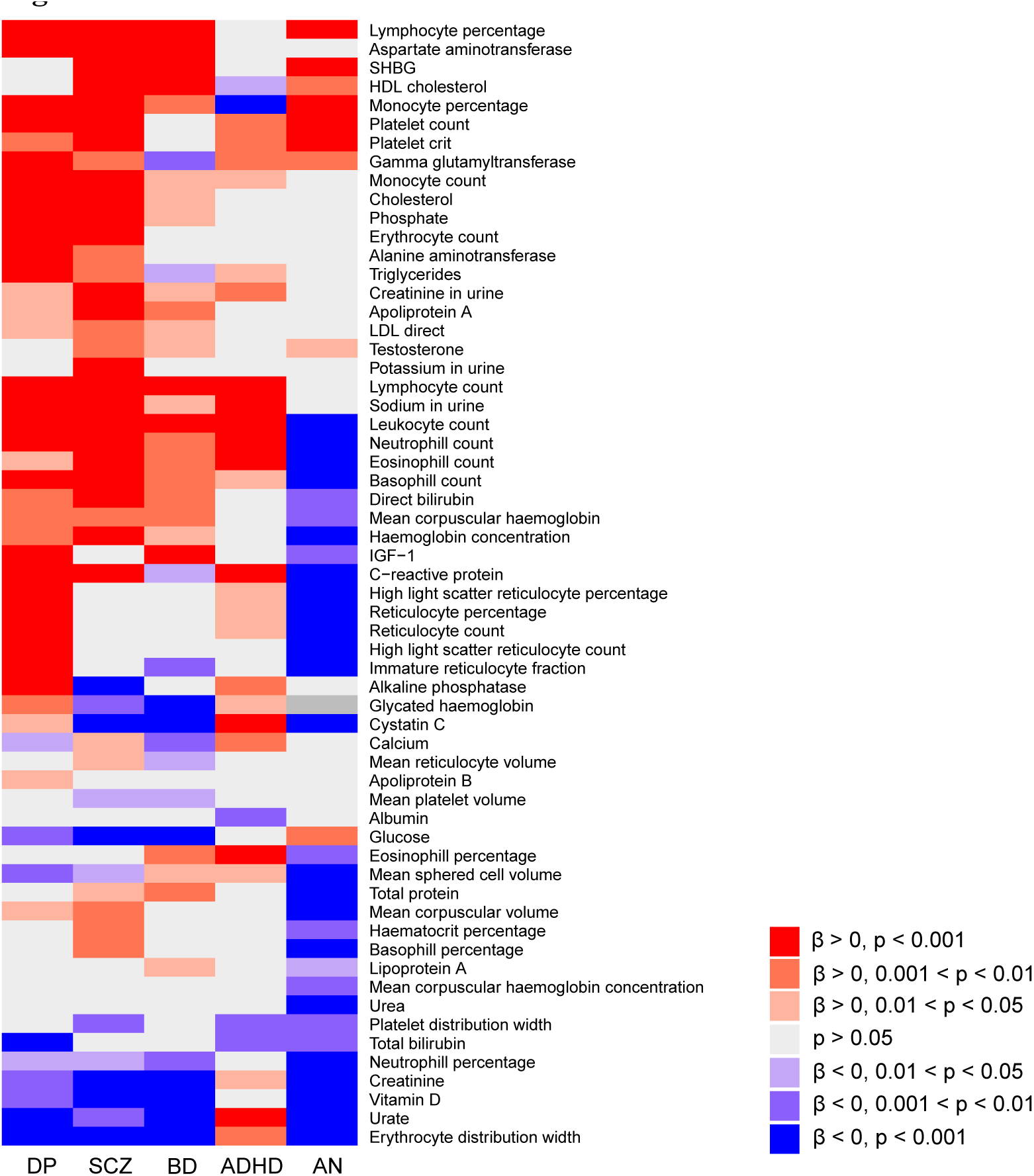
Causal effects of mental disorders on clinical traits. The heatmap depicts the causal effect of mental disorders (exposures) on clinical traits (mediators). Each column represents a mental disorder, while each row represents a clinical trait. The red bricks represent the promoting causal effects (β > 0). The blue bricks represent the inhibitory causal effects (β < 0).

**Figure S2.**
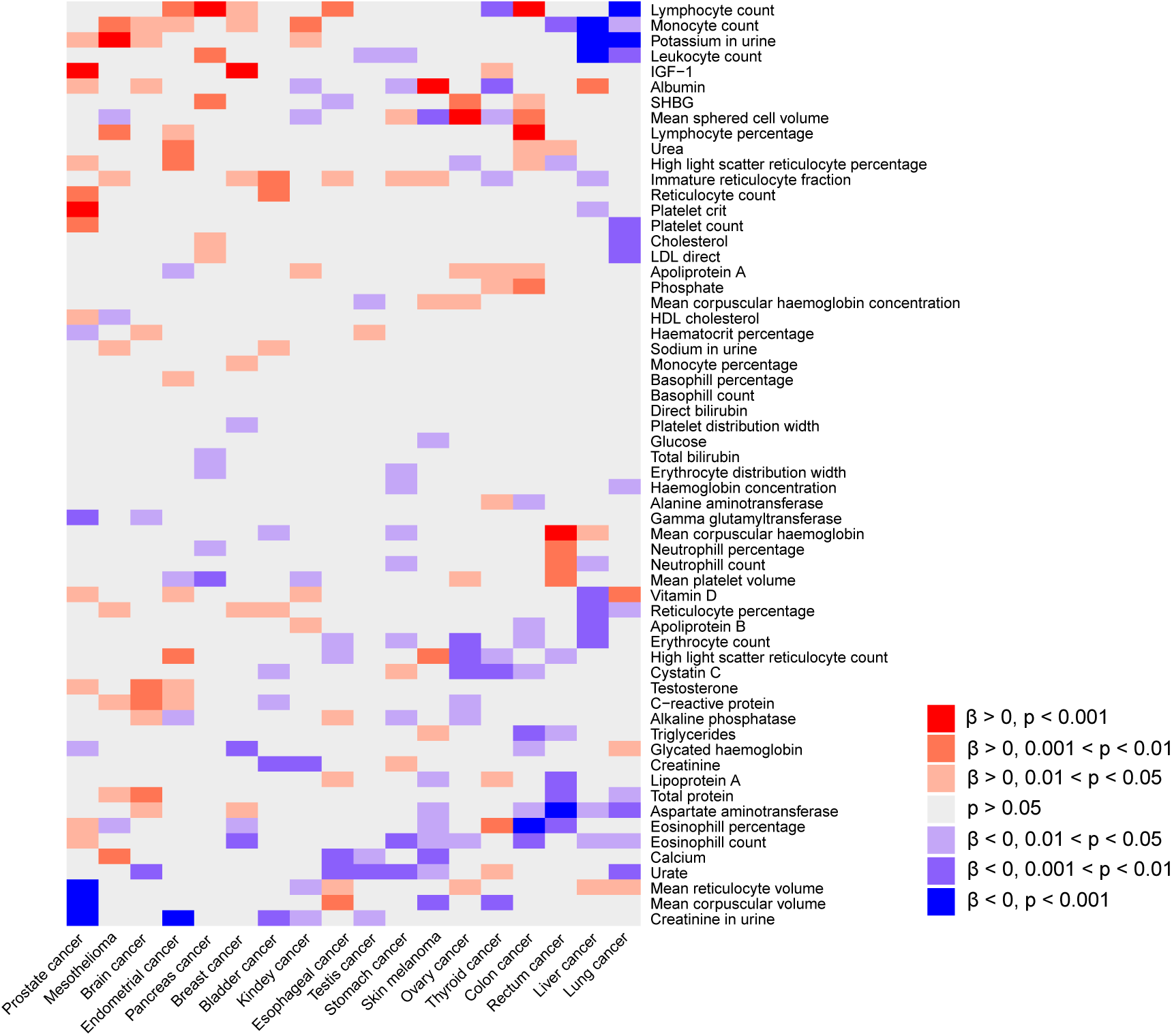
Causal effects of clinical traits on cancers. The heatmap depicts the causal effect of clinical traits (mediators) on cancers (outcomes). Each row represents a clinical trait, while each column represents a type of cancer. The red bricks represent the promoting causal effects (β > 0). The blue bricks represent the inhibitory causal effects (β < 0).

